# Percutaneous Nephrostomy Guidance by a Convolutional Neural Network Based Endoscopic Optical Coherence Tomography System

**DOI:** 10.1101/2024.02.06.24302404

**Authors:** Chen Wang, Paul Calle, Feng Yan, Qinghao Zhang, Kar-ming Fung, Chongle Pan, Qinggong Tang

## Abstract

Percutaneous nephrostomy (PCN) is a commonly used procedure for kidney surgeries. However, difficulties persist in precisely locating the PCN needle tip during its insertion into the kidney. Challenges for PCN needle guidance exist in two aspects: 1) Accurate tissue recognition, and 2) Renal blood vessel detection. In this study, we demonstrated an endoscopic optical coherence tomography (OCT) system for PCN needle guidance. Human kidney samples are utilized in the experiments. Different renal tissues including: 1) cortex, 2) medulla, 3) calyx, 4) fat, and 5) pelvis can be clearly distinguished based on their OCT imaging features. We conduct kidney perfusion experiments to mimic the renal blood flow. Our system can efficiently detect the blood flow in front of PCN needle using Doppler OCT function. To improve surgical guidance efficiency and alleviate the workload of radiologists, we employ convolutional neural network (CNN) methods to automate the procedure. Three CNN models including ResNet50, InceptionV3, and Xception were applied for tissue classification. All of them demonstrate promising prediction results, with InceptionV3 achieving the highest recognition accuracy of 99.6%. For automatic blood vessel detection, nnU-net was applied, and it exhibited intersection over unions (IoU) values of 0.8917 for blood vessel and 0.9916 for background.

## 1. Introduction

Urolithiasis is one of the most common urinary diseases with renal stones being a common manifestation. Over half a century, the prevalence of renal stones has exhibited a noteworthy increase: from merely 3.8% in the late 1970s, to the 8.8% in the late 2000s, and has arrived up to 11.0% in recent years [1, 2]. In renal stone surgeries, urinary diversion and entry of surgical tools are necessary for urolithiasis treatment procedures, including urine drainage, stone breaking, removing procedure (percutaneous nephrolithotomy, PCNL), and other therapeutic intervention [3]. Nevertheless, conventional access through the ureter is unavailable in cases of renal stone blockage or urinary tract infection. In response to this challenge, a novel percutaneous renal access method is proposed. Percutaneous nephrostomy (PCN) is a well-established and minimally invasive kidney access therapy, commonly employed in varied renal surgical procedures [4-6]. During that, a needle is inserted through patient’s back or flank, creating a path to the kidney [7]. Urine can be drained through this route. Following track dilation, surgical instruments can be introduced through this percutaneous pathway to effectively address nephrolithiasis [8]. The precise selection of the insertion position for the PCN needle is crucial to ensure its arrival at the renal collecting system inside the kidney. Following the principles of applied anatomy for percutaneous access, an optimal track needs to be short and straight. It should traverse the renal cortex tissue and medullary pyramid tissue, passing through the renal papilla into the calyx tip, ultimately reaching the renal pelvis [9]. A puncture that deviates from the specified tissue sequence poses a risk of complications. For instance, a needle insertion through the renal column between adjacent medullary pyramids may result in the tearing of calyx tissue, particularly after renal dilation. Additionally, an excessively long track may lead to renal vascular rupture [10]. Therefore, precise navigation of the needle is requisite for safe and effective PCN entry. Two major imaging modalities: ultrasound and fluoroscopy, along with additional techniques such as computed tomography (CT), robotic-assisted guiding and electromagnetic tracking, have been widely applied in guiding the needle during PCN procedures since it was invented [11-15]. The needle track can be visually acquired using these techniques, providing valuable information to assist physicians in their clinical judgment during renal access surgeries.

Although various conventional imaging techniques have been employed, PCN still encounters challenges due to the limited resolution of these imaging methods. Conventional imaging modalities typically provide an overall view of the kidney with needle track, but may lack the information required for local and precise needle positioning, as well as accurate tissue identification [16]. Single-puncture is critical in PCN since needle placement is an invasive procedure. Multiple punctures will increase the risk of various potential complications. Ultrasound, recognized for its safety and convenience, has been extensively employed in PCN needle guidance. However, recent studies on ultrasound-guided PCN have shown that the single-puncture success rate can range from 34.3% to 91.2% due to its limited visualization details [17-20]. Fluoroscopy is also an effective tool for renal access achievement due to its outstanding capacity in imaging contrast. Similar to ultrasound, however, its guiding performances are also not consistent among different research works, with the single-puncture success rates from 50.0% to 79.9% [20-23]. Additionally, CT and other assistive technologies are applicable in guidance improvement, but the single-puncture success rates are still expected to be improved [21, 22, 24]. Complications following PCN needle insertion can also cause suffering for patients. In previous percutaneous access studies, the reported overall complication rate exceeded 20% [25, 26]. Major complications associated with the percutaneous approach include severe hemorrhage [27], sepsis and septic shock [28], transgression of intra-abdominal viscus like colon, liver or spleen [29], pleural injury [30], etc. [31]. In addition, minor complications such as urine extravasation, hematuria and ureteric colic are more commonly observed in PCN procedures [29]. The occurrence of complications is primarily influenced by two main factors: 1) kidney tissue injury and 2) blood vessel rupture. Deviations from the correct tissue sequence during needle placement can lead to injuries in the renal pelvic tissue and other normal kidney structures, resulting in various challenges during surgeries. The issues include kidney displacement, collapse of collecting system, poor vision of renal bleeding situation, and renal stone fragments extravasation [9]. Damage to the blood vessels, especially on the segmental renal artery, can lead to hemorrhage, fistula, or segmental infarction. Severe hemorrhages which require transfusion, surgical exploration, and nephrectomy have been reported to occur in approximately 2% of patients undergoing percutaneous procedures. Particularly, the incidence rate tends to double in patients with coagulopathy [27, 32]. Furthermore, minor temporary postoperative bleeding has been reported to occur in approximately 95% of PCN cases [8]. Hence, there is a demand for a biomedical imaging device with improved needle tracking precision in PCN needle guidance.

To achieve enhanced accuracy in PCN needle puncture, various novel and improved methods based on conventional imaging modalities have been applied. For instance, combined utilization of ultrasound and fluoroscopy [33], adoption of transrectal ultrasonography probe [34], and cone-beam CT with live three-dimensional (3D) needle guidance [35] have been tested feasible in tracking the PCN needle. The incorporation of these techniques allows for the acquisition of high-quality overall imaging results of the entire kidney, thereby enhancing needle tracking and potentially reducing complication rates during PCN. However, the absence of precise local imaging ahead of the needle tip remains a limitation of accurate tissue recognition, impacting the safety and efficiency of the PCN needle placement. To address the issues related to tissue injury and blood vessel rupture, there is a critical need for an advanced imaging technique with higher imaging precision. Optical coherence tomography (OCT) is a well-established medical imaging modality with superior axial resolution at micrometer level, approximately over ten times higher than conventional imaging techniques like ultrasound, fluoroscopy, and CT [36-39]. It is capable of achieving subsurface tissue imaging with a penetration depth of several millimeters [40], so it is applicable for forward imaging of the tissue in front of the PCN needle tip during the puncture. To integrate the OCT into the PCN procedure, we designed a forward-view endoscopic OCT system which is available for imaging tissues ahead of the needle. The system combines an endoscope with OCT machine, transmitting imaging results from the distal end of the endoscope to the OCT scanner lens. Endoscope has been previously reported to assist PCN guidance [41, 42]. However, conventional endoscope has limitations for tissue recognition because it only provides the tissue surface vision without subsurface information. This means that it may not effectively predict the risk of needle tip damage to the underlying kidney tissue ahead of the PCN needle tip. Since our endoscopic OCT can acquire biological structures beneath the imaging surface, it has great potential in accurately identifying the tissue type in front of the PCN needle to miniaturize the risks of kidney tissue injury. Furthermore, it has been reported that endoscopic OCT system functions effectively in blood vessel detection, digestive tract imaging, and tumor margin analysis [43-45]. Doppler ultrasonography has been applied in PCN guidance access to depict the renal vasculature and avoid major blood vessel rupture [46, 47]. Doppler OCT, recognized for its effectiveness in imaging fluid flow, has been proved to be feasible in blood flow detection precisely [48, 49]. In our study, we implemented the Doppler OCT scanning function of our system to help observe the renal blood vessel ahead of the needle. This approach allows for the assessment of vascular size and blood flow speed, providing valuable information to evaluate the risks of renal blood vessel damage.

With the capabilities of tissue type identification and blood vessel detection, our OCT probe exhibits potential applications in PCN needle guidance. To further enhance surgical efficiency, we proposed the utilization of computer-aided methods to automate the image analysis procedure. Deep learning, particularly through the use of convolutional neural networks (CNN), has demonstrated significant utility in image analysis for feature extraction and transformation. CNN, within the realm of deep learning models, exhibits outstanding performance in tasks such as image classification and segmentation [50]. Therefore, CNN architectures have been widely employed in medical imaging research [51-53]. In recent years, CNN-aided image classification has found widespread application across various medical imaging modalities, including ultrasound [54], CT [55], fluorescence [56], X-ray [57], angiography [58], photoacoustic microscopy [59], etc. It has been clinically applied in the diagnosis of different diseases, intraoperative tissue assessment, and blood vessel segmentation. To assist endoscopic OCT imaging, our previous studies have illustrated the potential of using CNN methods in different applications, such as pig kidney tissue classification [60], epidural needle navigation [61], and Veress needle location [62]. Therefore, we continue to use CNN methods for human kidney tissue recognition in this research. Furthermore, deep learning has been applied in the image contents segmentation task [63]. nnU-Net is a deep learning based segmentation method which has outstanding performance in segmentations [64]. It has proven to be an effective tool in biomedical image analysis and quantification across different medical domains. Thus, nnU-Net has the potential to assist in the segmentation of renal blood vessel signals from the Doppler OCT imaging results in our research. In this study, we utilized the following CNN architectures to fulfill the two research objectives: 1) ResNet 50, Inception, and Xception for renal tissue recognition; and 2) an automatic pipeline nnU-Net for the blood vessel segmentation. A complete deep learning-aided OCT imaging platform was established to assist in needle guidance during PCN procedures.

## 2. Method

### 2.1 Endoscopic OCT system setup

To realize the subsurface imaging of kidney tissue ahead of the PCN needle, we built an endoscopic OCT system by using gradient-index (GRIN) rod lens as shown in Fig. 1(A). In the perpendicular direction to the optical axis, the refractive index decreases from the center to the edge to keep the light propagating inside the GRIN rod lens. The light path follows a sinusoidal path, with each complete sinusoidal path defined as one ‘pitch’. The length of the GRIN rod lens we used is an integral multiple of the pitch length. Thus, it can transmit the imaging information from one end to the other without any loss of spatial resolution. Typically, PCN needles used have a gauge size of 16 or 18, with length ranging from 120 to 200 mm [8, 65]. In our system, to accommodate the specifications of practical PCN needle, we utilized the GRIN rod lens with a diameter of 1.30 mm, and a length of 138.60 mm (equivalent to 2 pitches). Therefore, the GRIN lens we employed could fit the hollow bore of the PCN needle. The numerical aperture (NA) is 0.098 and the view angle is 11.0°.

**Figure 1.**
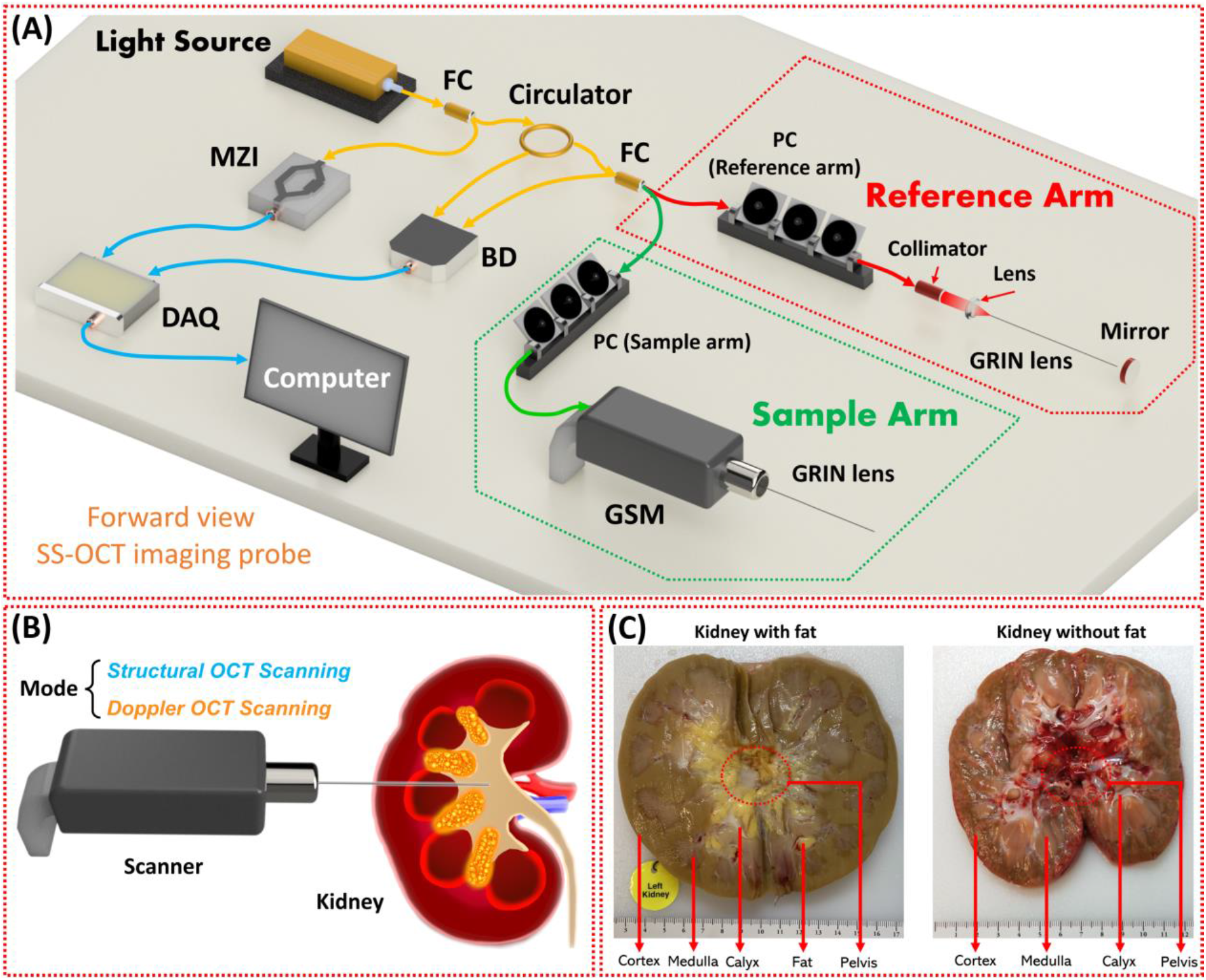
(A) Schematic of the forward-view endoscopic OCT imaging probe. (B) Examples of the human kidneys with and without renal fat. (C) Scanning procedure following the ideal PCN needle insertion.

A swept-source laser was used in our system as the OCT light source. It provides laser of around 25 mW output power, with a central wavelength of 1,300 nm and a tuning bandwidth of 100 nm. The axial-scan (A-scan) rate of the system can reach 200 kHz, enabling high-speed scanning and real-time OCT imaging. The output laser power was split in a 3%: 97% ratio using a fiber coupler (FC). The 3% portion was used to trigger OCT sampling through a Mach-Zehnder interferometer (MZI), and the remaining 97% was directed to an optical fiber circulator for OCT imaging. Since light can only travel in one direction within the circulator, the light input from any port can only exit from the next one. Consequently, the imaging laser input from port 1 exited the circulator through port 2, and then entered another FC. Subsequently, it was equally split into two parts: sample arm and reference arm. This configuration established a Michelson interferometer. In each arm, a polarization controller was incorporated to reduce noise during OCT imaging. Within the sample arm, the light was directed for imaging the renal tissue using a galvanometer scanning mirror (GSM), through a GRIN lens integrated in front of the scanning objective lens of the GSM. The proximal end surface of the GRIN lens was placed a little far away from the intermediate image plane of the GSM scanning objective lens. The backscattered signal which contained the tissue structural information returned to the FC for next processing. In the reference arm, a collimator transferred the light from optical fiber to space, and a convex lens converged the light into the GRIN lens. After entering the space, the light passed through another GRIN lens, which served to expand the optical length and compensate for light dispersion from the GRIN lens in the sample arm. Then, the light was reflected by a mirror and retraced its path back through the original route to the same FC where sample arm light returned. The two returned lights combined and the interference signals from the two arms were separated initially. Then, they are transmitted to a balanced detector (BD), which was utilized for common mode noise cancellation. Finally, the BD output signal propagated to a data acquisition (DAQ) board and a computer for processing and monitoring. This system has the highest scanning speed at 200 kHz with an axial scanning (A-Scan) resolution of 11 μm, a lateral scanning resolution of 20 μm, and a sensitivity of 92 dB. The extraordinary performance makes it available to achieve real-time imaging. (+structural and doppler scanning principles)

### 2.2. Experiments on human kidneys

This research was approved by the University of Oklahoma Institutional…… The organs are provided by Lifeshare of Oklahoma, and all the patient demographics were collected after obtaining consent. For our dataset, we utilized thirty-one human kidneys. Among them, seventeen were used for tissue recognition, seven were used for blood vessel detection, and the remaining seven were used for blind tests (five for tissue identification blind test and two for blood vessel detection blind tests).

#### 2.1.1. Structural imaging of different renal tissue

Structural OCT imaging can obtain biological structures of samples with very high resolutions. Hence, different renal tissues are potential to be distinguished based on their OCT imaging results. Seventeen human kidneys from thirteen donors (7 males and 6 females, with an average age of 53.15) were utilized in our experiments to test the feasibility of tissue recognition using our endoscopic OCT probe. To simulate a practical surgery situation, we first injected artificial urine into the renal pelvis through the ureter and tied the ureter end to prevent urine leakage. After that, we inserted our endoscopic OCT probe into the renal pelvis through the renal tissues. Considering the filled renal pelvis with urine as a liquid space, there would be a sudden decrease in backpressure from the renal tissues when our probe tip reached the renal pelvis. Structural OCT images of renal pelvis were acquired when the probe tip was inside the renal pelvis. Then, to obtain the OCT images of different renal tissues: cortex, medulla, calyx, and fat, we cut each kidney and exposed the mid pole. Different tissue types could be clearly distinguished based on their colors, shapes and positions as shown in Fig. 1(C). The cortex tissue constituted the outer brown layer of the kidney. Medulla tissue comprised a series of pyramid-like sections, distinguishable from the cortex by their different color. Calyces were the white tissue that surrounded the apex of the medulla pyramid units. Renal fat tissue was located between the adjacent medulla pyramids, identifiable by its yellow and soft features. We placed our probe tip on these different tissues and acquired their structural OCT images, separately. For each tissue type in every kidney, 10,000 cross-sectional OCT images were obtained for the next deep-learning work. It should be noted that in two kidneys, renal fat did not exist as shown in the example of Fig. 1(C).

#### 2.2.2 Doppler OCT imaging of renal blood flow

Doppler OCT is a technique assisting in detecting fluid flow detection. Therefore, it has the potential to visualize the renal blood flow in front of the GRIN lens in our endoscopic OCT probe. Seven human kidneys from six donors (1 male and 5 females, with an average age of 54.33) were utilized to detect the renal blood flow in the experiments using our endoscopic OCT probe. As shown in Fig. 2, the renal artery and vein were cannulated and perfused using a pulsatile perfusion pump. The perfusion solution we used included red blood cells (RBC) and Ringer’s solution [66]. The solution flow rate was set within the range of 100 −250 ml/min for normothermic *ex-vivo* kidney perfusion (NEVKP) [67]. A water bath heater was utilized to maintain the temperature of the perfusion solution at a constant 37°C, mimicking the normal human body temperature. Additionally, the ureter was cannulated with a tube connected to a beaker for urine collection. When the perfusion system started working, our OCT probe was inserted into the kidney being perfused. We fabricated a sharp and hollow needle which accommodated the GRIN lens and integrated the needle onto the GRIN lens for insertion. The OCT scanning was set to Doppler mode. We gradually reduced the insertion speed when Doppler signals of the flow were detected. Then, we started to record Doppler images with different distances between the OCT probe tip and the edge of the Doppler signals, which corresponded to the renal vascular walls. The schematic of the Doppler OCT imaging of renal blood flow is shown in Fig. 2(A), and the experimental setup is shown in Fig. 2(B).

**Figure 2.**
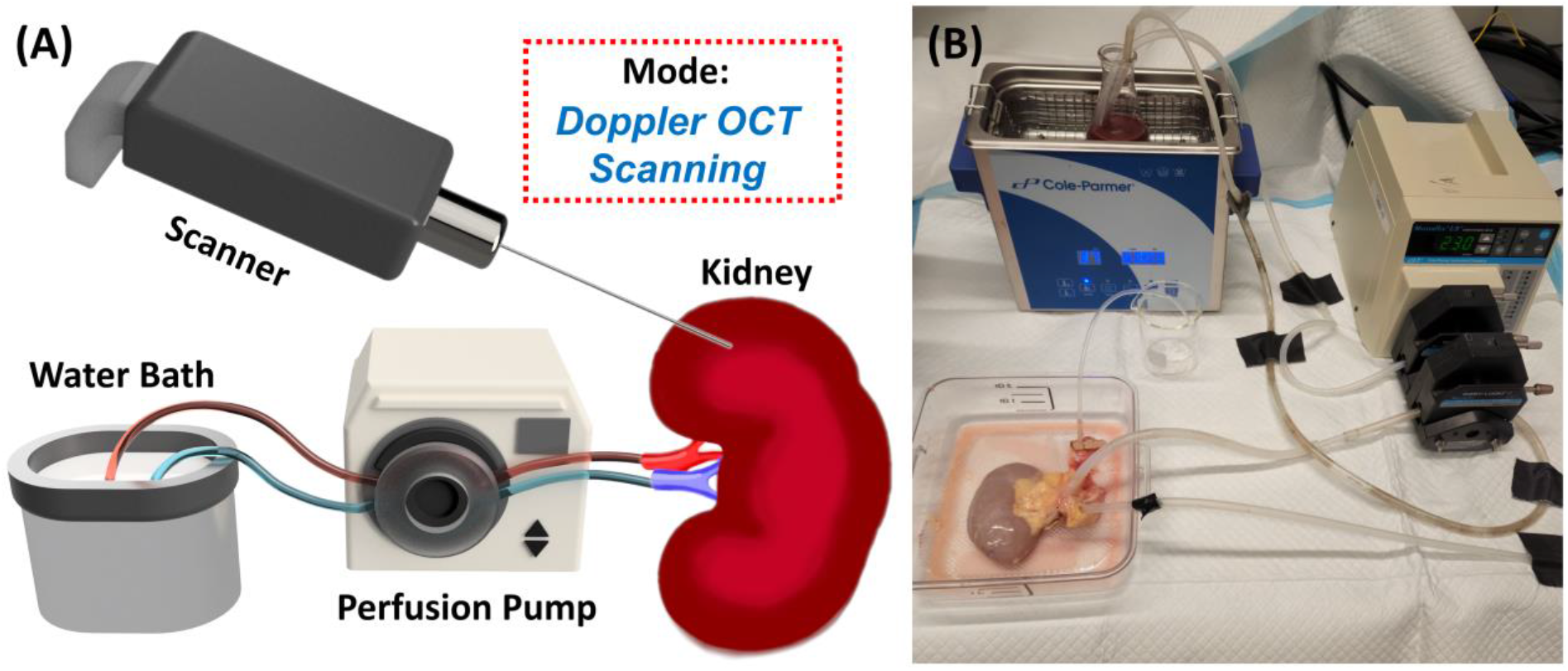
(A) Schematic of the perfusion pump system for Doppler OCT imaging of renal blood vessel. (B) Experimental setup of the perfusion system.

### 2.3. Deep learning method

In our study, we employed deep learning methods to enhance the effectiveness of tissue recognition and blood vessel detection during PCN procedure. CNN architectures were chosen for their widespread use in image processing, classification, and segmentation tasks. We used a Linux server with Ubuntu 20.04.6 LTS with 2 Nvidia RTX A6000 GPUs to process all the tasks.

#### 2.3.1 Renal tissue classification

CNN methods were utilized in automatic renal tissue classification and renal blood vessel detection. In our study, we selected three architectures: ResNet50, Inception, and Xception, all of which have been applied in medical imaging classifications areas, such as pneumonia X-ray [68], breast cancer histopathology [69], and diabetic retinopathy [70]. In our model, 12 kidneys were used for cross-validation and 5 kidneys were used for testing as shown in Fig. 3(A). 10,000 OCT images for each tissue type in each kidney were utilized in the model training.

**Figure 3.**
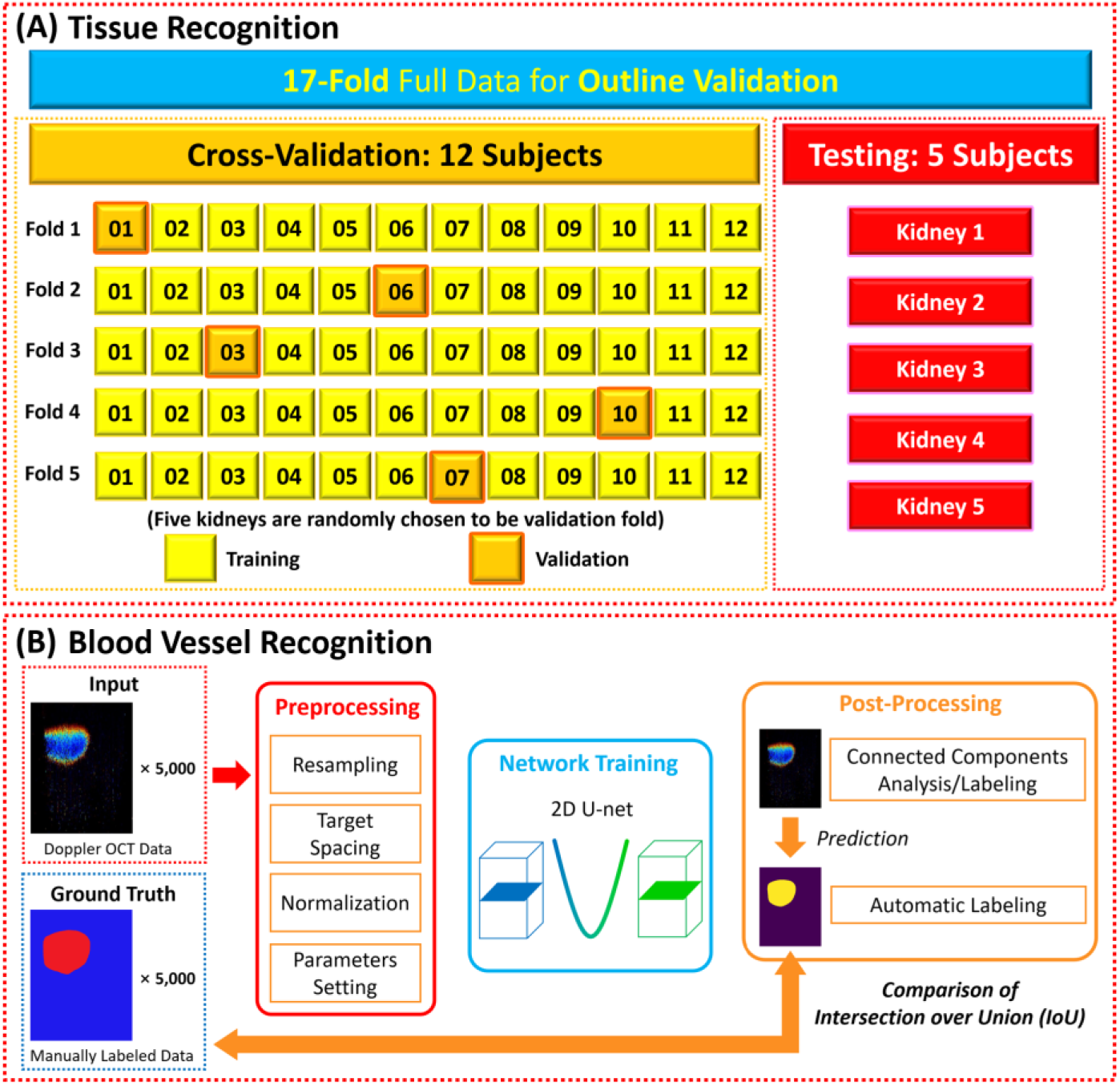
(A) Cross-validation and cross-testing methods for renal tissue recognition using CNN. (B) Procedure of using nnU-Net for renal blood vessel segmentation.

#### 2.3.2. Renal blood vessel segmentation

In total, we acquired 5,000 Doppler OCT images from all the seven perfused kidneys. Then we manually labeled the blood vessel areas on each image based on the Doppler signals. The labeled results were considered as the gold standard. U-net is a convolutional network widely used in biomedical image segmentation applications [71]. nnU-Net is an improved version of U-Net [64]. We applied 2D U-net configuration option of nnU-Net, and the procedure is shown in Fig. 3(B). We set the model with 1,000 epochs, and the stochastic gradient descent with a high initial learning rate of 0.01 and the large Nesterov momentum of 0.99. Intersection over union (IoU) was used as the segmentation metric.

### 2.4 Blind tests of the CNN models

To mimic the practical conditions and further validate the feasibility of our system, we further did blind tests on new kidneys. Five extra kidneys were used to test the tissue recognition. We followed the same method used in the previous imaging. For those five kidneys, we did the blind tests of cortex, medulla, calyx, fat, and pelvis on them, separately. For instance, to the first kidney, we inserted our probe into the kidney and started the OCT scanning. When we found the OCT imaging result looked like the cortex, we stopped insertion and recorded the results. After image recording, we took out our probe and inserted a short stainless-steel tube with the same size as the GRIN lens to the same depth of the OCT probe insertion. Thus, the tip of the stainless-steel tube corresponded to the OCT imaging position as shown in Fig. 4(A). We did the same experiments on the five kidneys for the five tissue types. After all the experiments ended, we cut the imaging position of the tissue together with the tube and confirmed that all the tissues followed the types we visually recognized. For each tested kidney, we obtained 1,000 images for testing.

**Figure 4.**
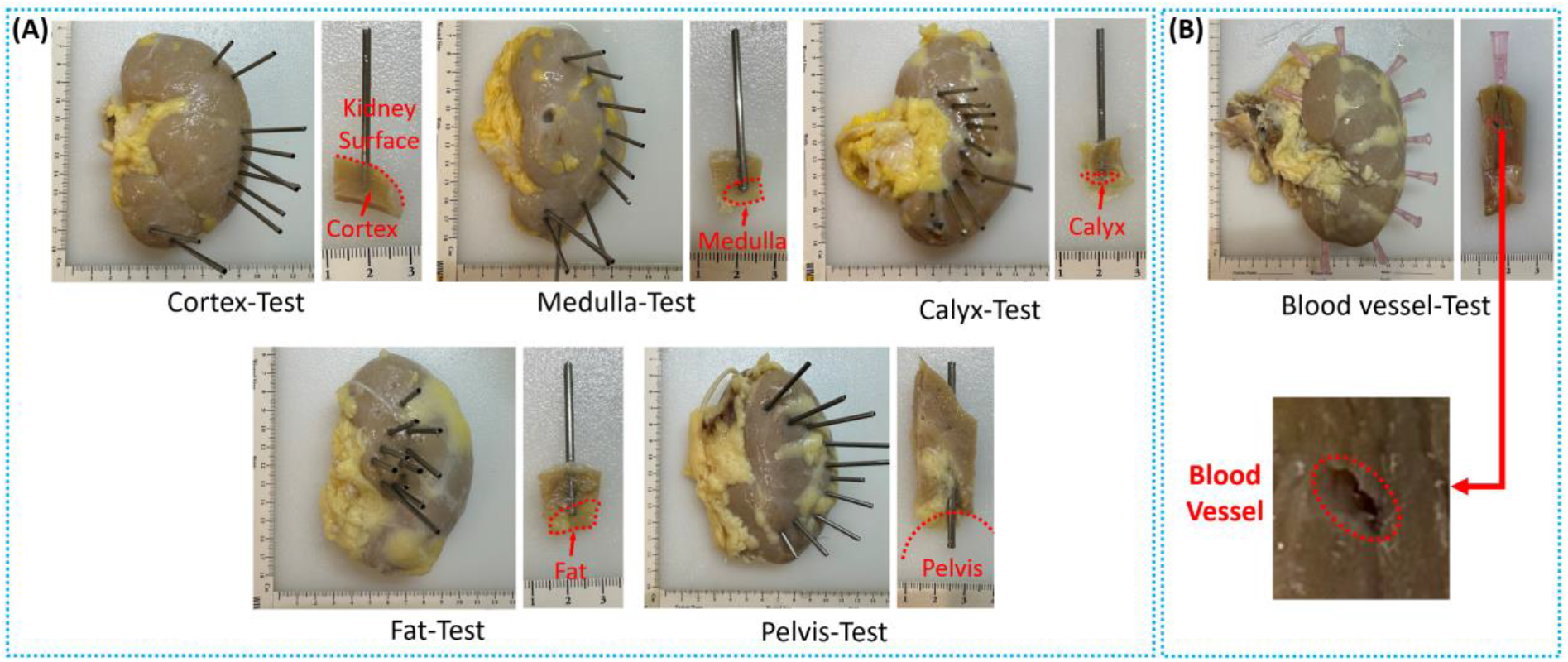
(A) Pictures of the five kidneys for tissue recognition test. (B) Pictures of the kidneys for renal blood vessel detection test.

Two extra kidneys (from two female donors with an average age of 67.5) were used in blood vessel detection. Same to the previous blood vessel detection experiments, we perfused blood solution through the renal artery and vein and cannulated the ureter of each kidney. Hollow tubes were applied on the GRIN lens to achieve the puncture into whole kidneys. Ten insertions were executed on each kidney, and 100 Doppler OCT images were acquired for testing. The tissues that the probe was inserted through were cut with the tube as shown in Fig. 4(B). All the 100 images with blood vessel signals were from the insertions that renal blood vessels were found on the route.

## 3. Results

### 3.1. OCT imaging results

#### 3.1.1. Renal tissue recognition results

To realize the accurate PCN needle location, our endoscopic OCT probe was utilized to image different renal tissue positions including: 1) cortex, 2) medulla, 3) calyx, 4) fat, and 5) pelvis. OCT structural imaging results of these different renal tissues were illustrated in Fig. 5. The imaging positions were demonstrated in Fig. 5(A) and the corresponding OCT imaging results were shown in Fig. 5 (B). The GRIN lens tip was positioned on the renal tissues, and the scanning procedure was initiated after that. Three-dimensional (3D) results were acquired, and the two-dimensional (2D) cross-sectional images were extracted from them. According to the results, different human renal tissues exhibited separate OCT imaging features. The cortex and medulla presented relatively homogeneous pixel distributions in the tissue areas, with a gradual overall decrease in pixel values in the vertical direction. However, the imaging depth of medulla was much larger than that of the cortex. Calyx tissue, formed by mucous membranes covered by connective tissue, showed loose-to-dense biological features, resulting in transverse dark-bright stripes in the OCT images based on the intensity changes. Renal sinus fat, composed of adipose tissue containing large globules of fat (lipid droplets) and fiber structural networks, was identifiable in OCT images through its bright spots and reticulate patterns. The pelvis, a space that collects urine, did not show any tissue ahead when our OCT probe tip arrived. It was featured by its isolated bright line on the top which is the surface of the GRIN lens, and the space without tissue signal under it.

**Figure 5.**
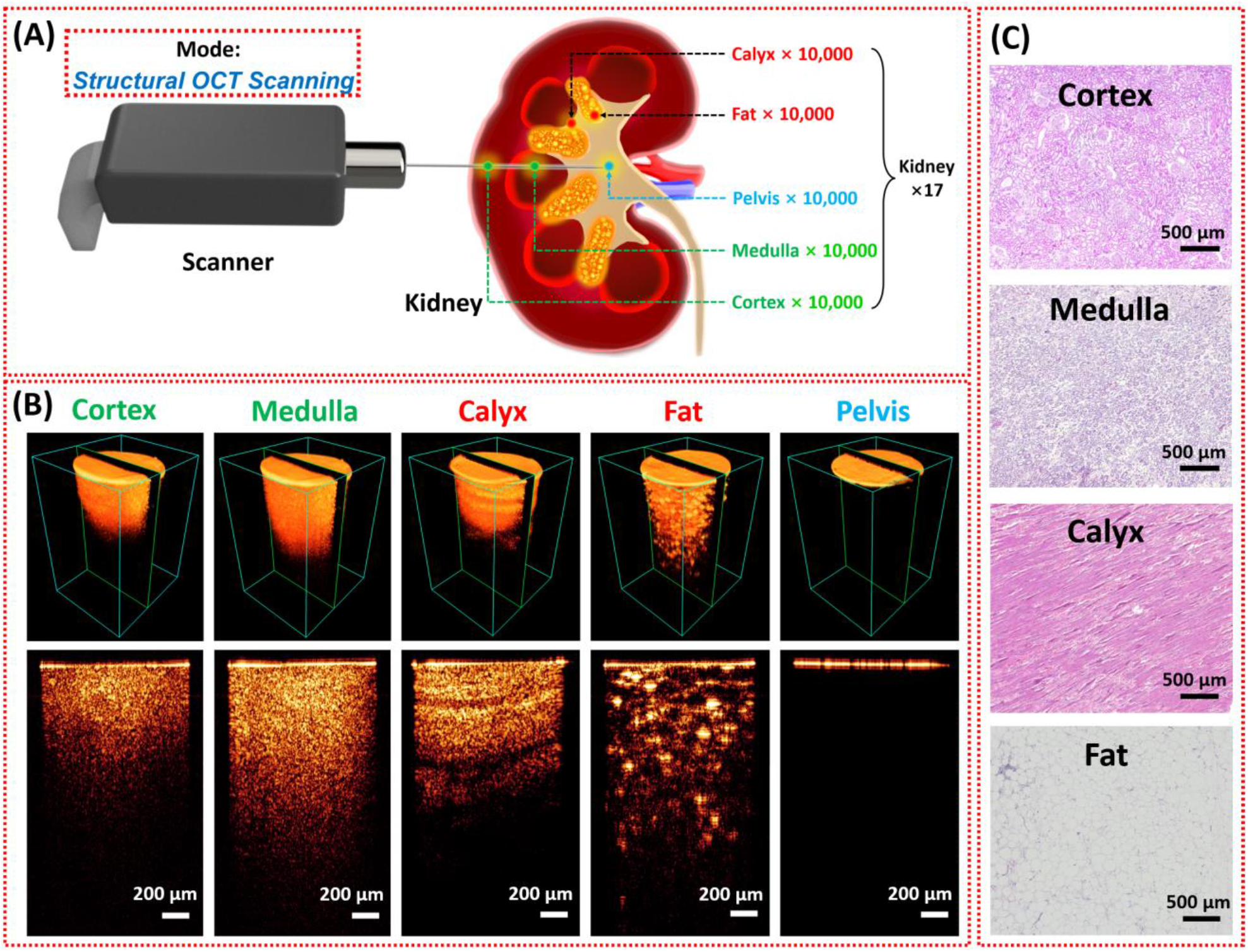
OCT imaging results. (A) Imaging procedure by using our OCT probe. (B) Imaging results of different renal tissue types. (C) Corresponding histology results of different renal tissue types.

#### 3.1.2 Renal blood vessel segmentation results

Three groups of representative Doppler OCT imaging results of renal blood vessels with different sizes were shown in Fig. 7. The green lines on the top corresponded to the positions of the GRIN lens surface cross sections. The blood flow velocity in different positions could be distinguished through the phase-based colors. In each group, the procedure of our OCT probe getting closer to the renal blood vessels were demonstrated in sequence. It can be seen from the OCT images of each group that when the blood vessel was closer to the probe, its cross-sectional shape was flattened due to the pressure. From the results, the blood flow in front of the probe could be efficiently detected. Therefore, our OCT probe is potential to help the physicians know the blood vessel sizes and its distance to the needle tip, assisting them to estimate the risk of the blood vessel injury and determine following steps to guarantee the surgery safety and efficiency. As listed in Fig. 7, renal blood vessels with different sizes were demonstrated. As the needle insertion proceeded, the distances between the blood vessel and the GRIN lens surface (green dashed lines) became closer, and the shape of the blood vessel cross sections were pressed. The renal blood vessels with < 200μm could be clearly detected. Therefore, our OCT endoscope system can effectively assist the anesthesiologist in avoiding the puncture of renal blood vessels and mitigating the risk of bleeding.

### 3.2. Deep learning prediction results

#### 3.2.1 Prediction of tissue recognition

Deep learning results for the tissue recognition were shown as follows. Three CNN architectures: ResNet50, InceptionV3, Xception were applied. The average confusion matrix of these three models shown in Table 1 illustrated the accuracy values for each of the five validation folds (one kidney for each fold) in the cross-validation part (12 kidneys used). Very promising prediction results were achieved from the confusion matrix. Among three architectures, InceptionV3 performed the best prediction, with 99.6% of average accuracy. ResNet50 and Xception also showed high prediction accuracies of 98.9% and 99.5% respectively. Table. 2 demonstrated the accuracy values of the cross-validation results from the three CNN models, respectively. The pelvis provided an ideal cross-validation prediction, with no true pelvis predicted to other tissues and no other tissues were predicted to pelvis. This should be attributed to the obvious space under the GRIN lens surface in the OCT imaging results. The calyx also exhibited very promising predictions, with occasional instances of other tissues such as cortex and fat being predicted as calyx. The fat had the relatively lowest prediction results, and the wrong predictions focused on cortex and calyx. The micro-average receiver operating characteristic (ROC) curves among the five tested kidneys were included in Fig. 6, with all of the curves superimposed together and the areas under the curve (AUC) reaching 1.00.

**Table 1.** Accuracy values of the cross-validation results in different CNN architectures.

|  | Fold 1 | Fold 2 | Fold 3 | Fold 4 | Fold 5 | Average | Std Error |
| --- | --- | --- | --- | --- | --- | --- | --- |
| **ResNet50** | 99.2% | 99.8% | 98.2% | 98.7% | 98.7% | **98.9%** | **0.27%** |
| **InceptionV3** | 99.7% | 100.0% | 99.3% | 98.9% | 99.9% | **99.6%** | **0.21%** |
| **Xception** | 100.0% | 99.9% | 98.6% | 99.0% | 99.9% | **99.5%** | **0.29%** |

**Table 2.** Average confusion matrix of all three CNN architectures.

|  |  | Predicted |  |  |  |  |
| --- | --- | --- | --- | --- | --- | --- |
|  |  | Cortex | Medulla | Calyx | Fat | Pelvis |
| Truth | Cortex | **9989.4 ± 10.6** | 0.0 ± 0.0 | 10.6 ± 10.6 | 0.0 ± 0.0 | 0.0 ± 0.0 |
|  | Medulla | 0.0 ± 0.0 | **9998.0 ± 2.0** | 2.0 ± 2.0 | 0.0 ± 0.0 | 0.0 ± 0.0 |
|  | Calyx | 0.0 ± 0.0 | 0.0 ± 0.0 | **9999.8 ± 0.2** | 0.2 ± 0.2 | 0.0 ± 0.0 |
|  | Fat | 9.4 ± 9.4 | 0.0 ± 0.0 | 10.8 ± 10.8 | **9979.8 ± 20.2** | 0.0 ± 0.0 |
|  | Pelvis | 0.0 ± 0.0 | 0.0 ± 0.0 | 0.0 ± 0.0 | 0.0 ± 0.0 | **10000.0 ± 0.0** |

**Table 3.** Confusion matrix of the tissue recognition blind test.

|  |  | Prediction |  |  |  |  |
| --- | --- | --- | --- | --- | --- | --- |
|  |  | Cortex | Medulla | Calyx | Fat | Pelvis |
| Testing Sample | P1 | 986 | 0 | 0 | 14 | 0 |
|  | P2 | 2 | 998 | 0 | 0 | 0 |
|  | P3 | 0 | 0 | 916 | 84 | 0 |
|  | P4 | 31 | 0 | 0 | 969 | 0 |
|  | P5 | 0 | 0 | 0 | 0 | 1000 |

**Figure 6.**
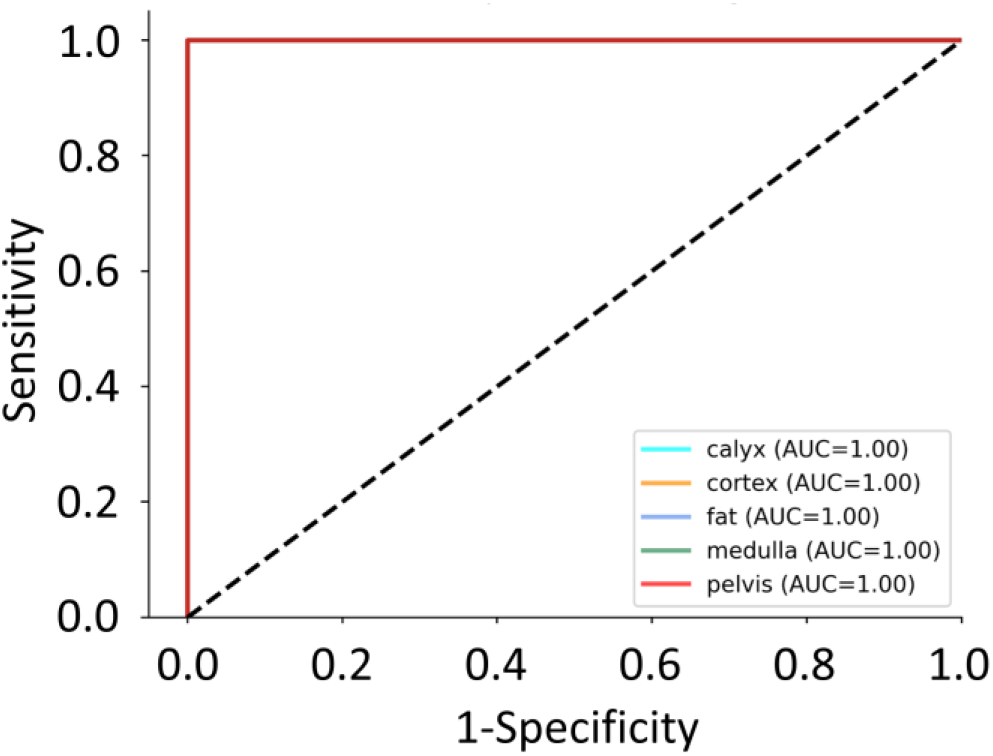
Micro-average ROC curve for five tested kidneys.

#### 3.2.2 Segmentation of renal blood vessels

In our study, nnU-Net was applied for renal blood vessel segmentations. Five thousand Doppler OCT imaging results were applied in training the model. To distinguish blood vessel signal from the background, we manually labeled the blood vessel signals in red and background in blue as shown in Fig. 7(A). The manually labeled results were considered as the ground truth of the segmentation data.

**Figure 7.**
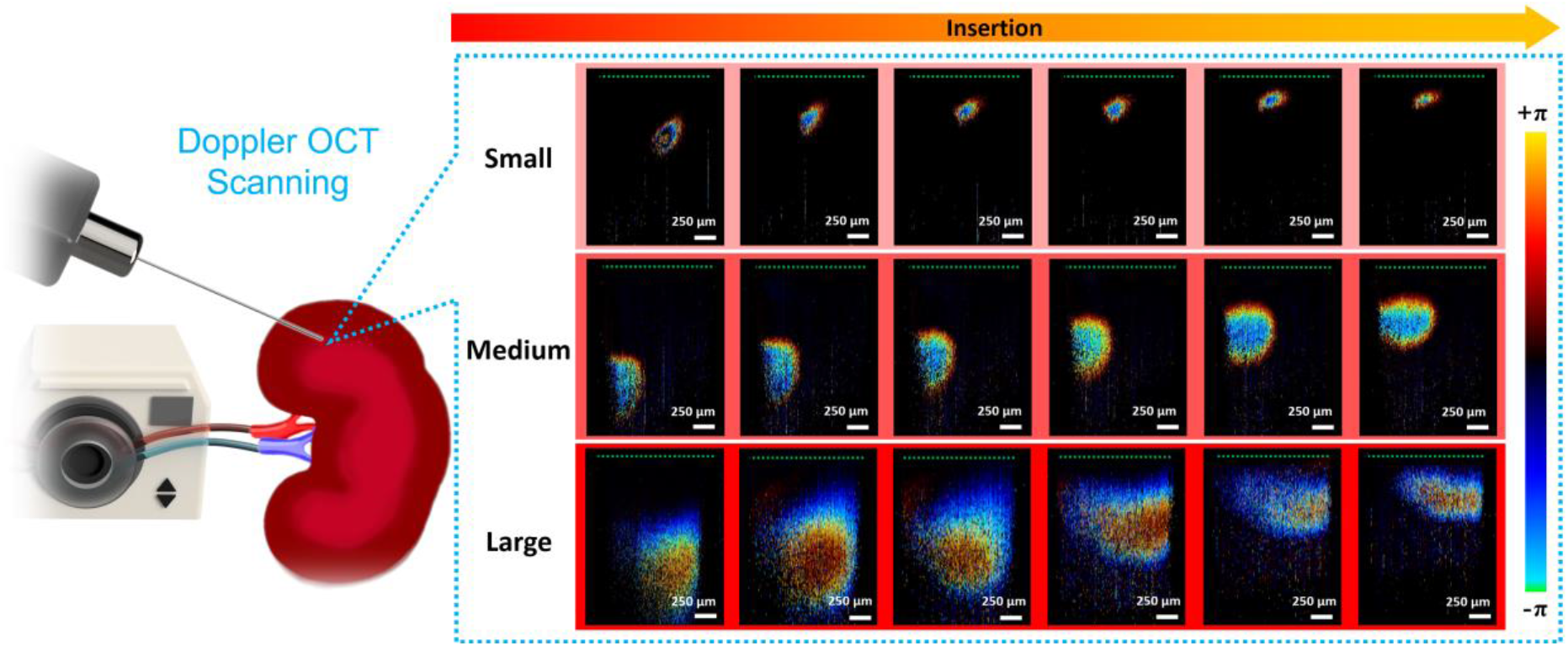
Doppler imaging results of blood flow detection with the renal blood vessels in different sizes.

**Figure 7.**
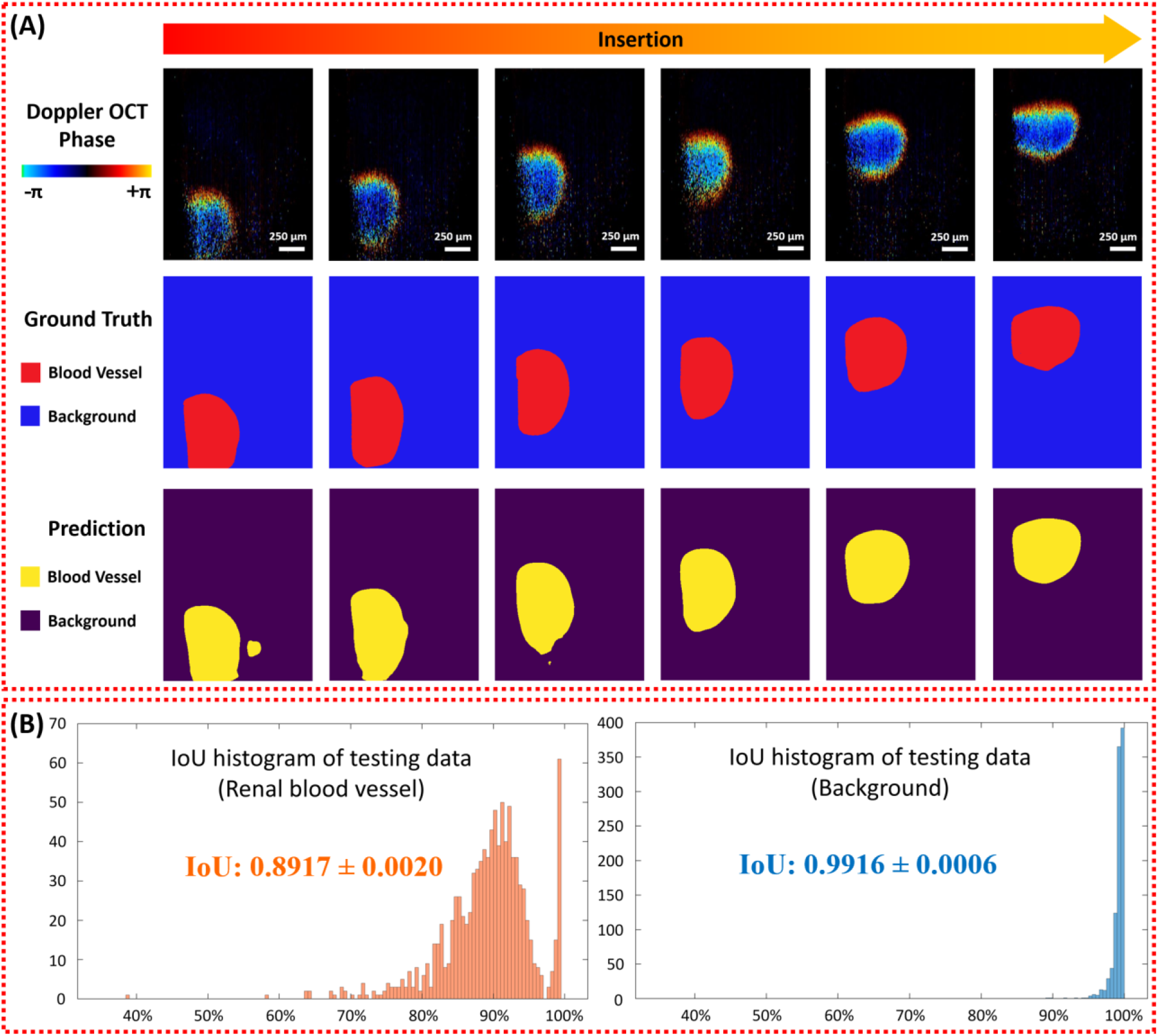
(A) Segmentation of renal blood vessel by using nnU-Net. (B) Histograms for IoU blood vessel for test dataset.

The nnU-Net model performed a cross-validation of five folds. For prediction of test results, the library used an ensemble model of the 5 folds. Fig. 7(B) shows the results for the test dataset. Intersection over union (IoU) was used as the standard to evaluate the prediction results. We obtained an optimistic average value 0.8917 for renal blood vessel detection, and 0.9916 for background segmentation.

### 3.3 Prediction results of blind tests

Blind insertion tests were conducted on five kidneys, with 1,000 images acquired for each kidney. The confusion matrix of the blind test was shown in Fig. 8(A). The average prediction accuracy among all the samples reached 97.38%. Same to the training model, pelvis performed the ideal prediction with no false positive or false negative predictions. Cortex and medulla also provided very promising prediction with over 98% accuracies. However, the prediction of calyx was not very promising, with 0.84% of the calyx data were recognized as fat. This might be because in kidney anatomy, renal fat usually existed beside the calyx, and they sometimes showed up in same OCT imaging results.

**Figure 8.**
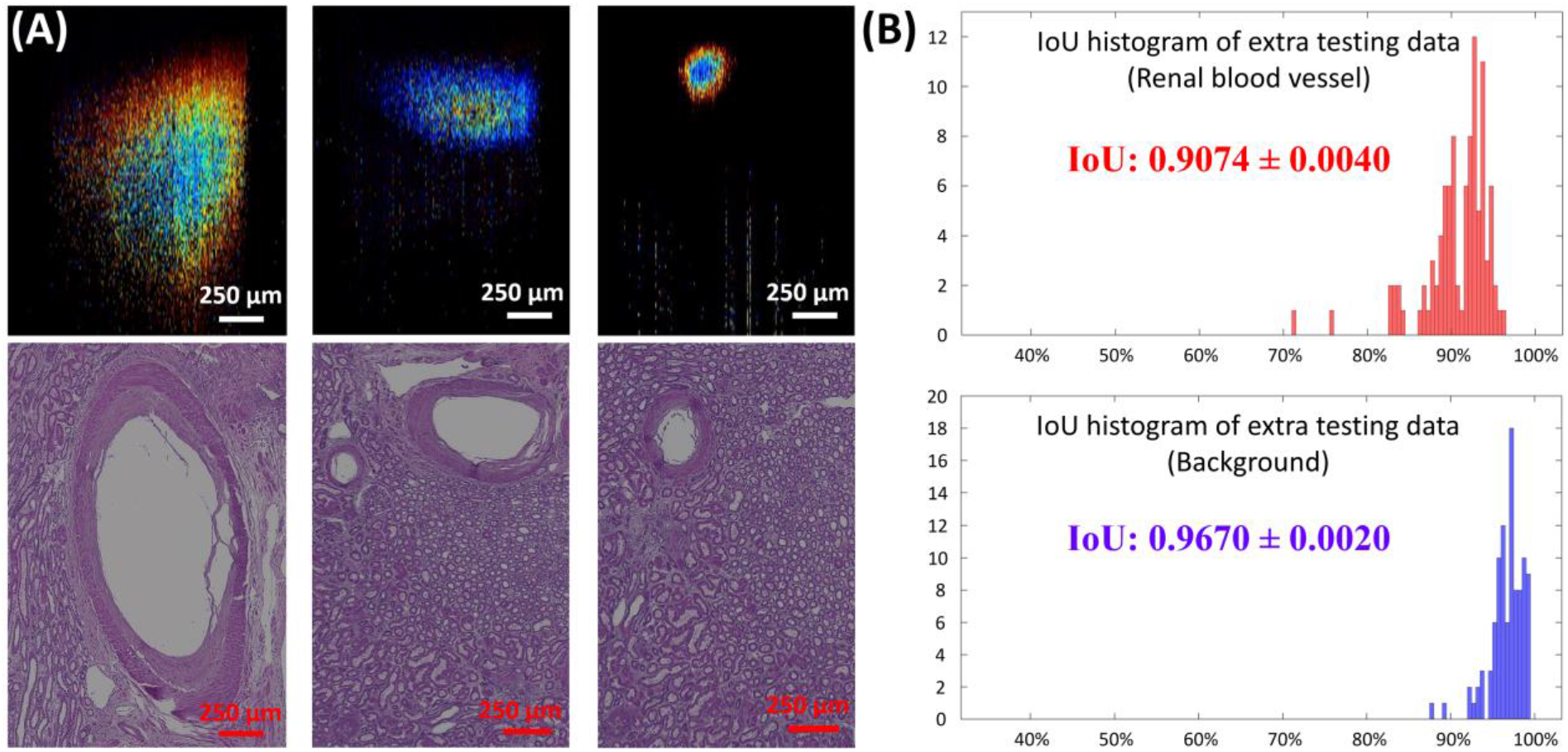
(A) Doppler OCT images of detected renal blood vessels and the corresponding. (B) Histograms for IoU blood vessels for the extra 100 test dataset.

The blind segmentations of renal blood vessels results were showed in Fig. 8(A). Three examples of the Doppler OCT imaging results with different blood vessel sizes were demonstrated. To further perform the reliability, we conducted the histology of the corresponding imaged blood vessels as demonstrated in Fig. 8(B), and the results were also illustrated. It is clear that the Doppler signals could correspond to the blood vessel structures from histology pictures. The IoUs of both blood vessel and background were over 0.90, which proved the promising performance of the blind tests.

## 5. Discussion

PCN is a commonly employed surgical procedure for accessing the kidney for various treatments. However, the PCN needle insertion demands more precise imaging guidance to improve safety and efficiency. In this study, we investigated an endoscopic OCT probe with a forward-viewing capacity in front of GRIN lens. The GRIN lens we utilized in our probe system is designed to fit inside the hollow bore of the PCN needle. This allows for OCT scanning of the needle tip position during the needle insertion process without introducing any extra invasiveness. Human kidneys were applied in the experiments to test of our system. To ensure efficient and safe needle tip location, we conducted tests on tissue recognition and blood vessel segmentation functions. Furthermore, deep learning methods were applied in this research to automate the image processing and identification for the purpose of enhancing the surgical guidance efficiency and alleviating the workload of physicians. CNN, as a popular method widely used in image classification and segmentation, was employed in our study.

Regarding tissue recognition, OCT images of five different renal tissue types including: 1) cortex, 2) medulla, 3) calyx, 4) fat, and 5) pelvis were scanned using our OCT probe. These tissues exhibited varying imaging characteristics in the OCT results, offering potential features for distinguishing. Physicians are able to ascertain the type of tissue in front of the PCN needle tip, aiding in decisions on whether to continue, halt, or reschedule the needle insertion. CNN architectures including ResNet50, InceptionV3, and Xception were used for automatic renal tissue recognition. In total, seventeen human kidneys were utilized, with twelve of them allocated for model training and the remaining five for testing. The average cross-validation confusion matrix of the three architectures showed very promising results. It revealed that there was no mistake in the prediction of pelvis. Considering that the pelvis serves as the target for PCN needle insertion, this result significantly demonstrated the reliability of our OCT probe. Additionally, the predictions for other renal tissues also exhibited exceptionally high accuracy rates from cross-validation results. The medulla and calyx attained 99.980% and 99.998% average accuracies, respectively. Prediction rates for cortex and fat were slightly lower, reaching 99.894% and 99.798%, respectively. For cross validation results, all three CNN architectures showed over 98% accuracies for every fold. Among them, InceptionV3 presented the most favorable prediction performance with a 99.6% accuracy rate, while ResNet50 achieved 98.9%, and Xception reached 99.5%. Slight variations in predictions across different folds were observed, and this phenomenon could be attributed to the physiological differences among the five tested kidney samples. Moreover, the results from an extra blind test further revealed the feasibility of tissue recognition. Similarly, there were no errors in predicting calyx, and the performances of other tissues were also favorable (>96.9%). However, the prediction of calyx was less promising at 91.6%. This could be attributed to kidney anatomy, where renal fat is typically situated beside the calyx, and fat tissue sometimes appeared in the calyx OCT images. It is noted that the kidney sample we used for blind tests happened to have a considerable amount of fat. From the blind test confusion matrix, all incorrect predictions for the calyx were identified as fat. To prevent blood vessel ruptures, human kidneys were perfused to simulate a situation where the kidneys remained alive. Doppler OCT signals, corresponding to renal blood flow, were obtained when blood flow was present in front of the OCT probe tip during scanning. The signal from the cross sections of blood vessels could be clearly discerned from the background, validating the feasibility of blood vessel detection using our system. For automatic blood vessel segmentation, the nnU-Net architecture was applied since it was a model configured for segmentation. In the experiments involving five perfused kidneys, five thousand Doppler OCT images with blood vessel signals were obtained. IoU was utilized to evaluate the blood vessel detection performance. The IoU level of renal blood flow signal was 89.17%, and that of the background signal reached 99.16%. Extra tests on two other kidneys were conducted to further substantiate the feasibility of blood vessel detection, revealing IoU levels of 90.74% for blood flow and 96.70% for background, respectively.

Since real-time feedback is significant for PCN needle guidance, the speed of tissue recognition and blood vessel detection is of great importance. We used a Linux server with Ubuntu 20.04.6 LTS that has 2 Nvidia RTX A6000 GPUs for all data processing in CNN tasks. For tissue recognition, predicting 1,000 images with a batch size of 32 performed an average prediction time of 6.07 ms per image. For blood vessel segmentation, the average processing time was 250 ms. These results affirmed the real-time performance of our OCT imaging platform.

In the future, we aim to enhance the efficiency of our OCT platform through system upgrades and CNN model optimization. On one hand, we plan to use polarization-sensitive OCT (PS-OCT) to further improve the tissue classification accuracy. PS-OCT signifies an improvement upon traditional intensity OCT, offering polarization-related tissue information through the effects of diattenuation and birefringence [72]. Hence, PS-OCT has the potential to improve the tissue prediction accuracy. On the other hand, we will conduct experiments using *in-vivo* animal models to further test the feasibility of our system. To further improve tissue recognition and blood vessel detection, we plan to improve our CNN models and upgrade the computer to improve the processing efficiency.

## Data Availability

All data produced in the present study are available upon reasonable request to the authors.

